# Differentiating central nervous system demyelinating disorders using Graph Attention Networks

**DOI:** 10.64898/2025.12.29.25343068

**Authors:** Stamatis Choudalakis, Anna Rapti, Dimitris Karathanasis, George A. Kastis, Maria-Eleftheria Evangelopoulos, Clio P. Mavragani, Nikolaos Dikaios

**Affiliations:** Department of Physiology, School of Medicine, National and Kapodistrian University of Athens, Athens, Greece; Mathematics Research Center, Academy of Athens, 4, Soranou Efesiou str., 11527 Athens, Greece; First Department of Neurology, National and Kapodistrian University of Athens, Aeginition Hospital, Athens, Greece

**Keywords:** semi-supervised clustering, graph neural networks, multiple sclerosis

## Abstract

Autoimmune demyelinating central nervous system (CNS) disorders encompass a wide array of clinical entities ranging from the organ specific Multiple Sclerosis (MS) to systemic autoimmune diseases (SADs) such as Systemic Lupus Erythematosus (SLE) and Sjögren’s syndrome (SS). Despite international research efforts, distinction of these entities at clinical, imaging and laboratory level remains challenging, with almost 20% of patients being misdiagnosed with MS, out of which more than 50% carry the misdiagnosis for at least 3 years, while 5% are misdiagnosed for 20 years or more. This work aims to identify early biomarkers that can discriminate MS from other clinical entities that are MS mimickers. For this reason, we have meticulously curated a unique biobank with serum, DNA, RNA, and cerebrospinal fluid (CSF) samples from 396 treatment-naïve patients who presented with a demyelinating episode and for which we have recorded over 300 clinical, serological, and imaging parameters. These patients have undergone follow up at 6 months and 12 months from their first demyelinating episodes and have been categorised as follows: MS, SAD with CNS involvement, and demyelination with autoimmune features (DAF). A hybrid model for semi-supervised tabular classification is proposed that integrates a graph attention network with dynamic graph learning via Random Forest proximity and k-NN graphs with tabular prior-data fitted network for direct classification. The model also performed feature selection, self-supervised contrastive learning, self-training and data augmentation.

## 1 Introduction

Multiple sclerosis (MS) affects more than 2.8 million individuals worldwide [1] and even though it is the most common central nervous system (CNS) demyelinating disorder, several clinical entities are major MS mimickers and need to be taken into consideration during differential diagnosis and therapy, especially after the revision of the McDonald criteria in 2017 [2–5]. The primary objective of this project is to discriminate the intricate underlying mechanisms of multiple sclerosis (MS) and autoimmune disorders with central nervous system (CNS) involvement that present overlapping clinical manifestations.

To that end, this work integrated transcriptomic, proteomic, and immunophenotyping data derived from peripheral blood and cerebrospinal fluid (CSF) analyses and corroborated these findings with clinical, imaging, and laboratory data specific to MS and CNS autoimmune patient cohorts. A unique patient cohort has been established, of 396 patients diagnosed with MS spectrum and CNS autoimmune disorders at 12 months after their initial demyelinating episode [6]. Based on a recent publication [6], 1/4 of patients who experience a first demyelinating episode with initial suspicion of MS, are eventually categorized in the CNS autoimmune group. Of the latter, 3/4 belong to the recently described clinical entity “Demyelination with autoimmune features” (DAF), whose pathogenetic mechanism is currently unknown, and the optimal therapeutic intervention unstudied. This group of patients further highlights the urgency of developing biomarkers that have the potential to differentiate patients with SAD and CNS involvement from those with DAF, or MS, and lead to the discovery of new pathogenetic pathways and thus the development of appropriate therapeutic strategies for these patients.

In view of these diagnostic and therapeutic challenges, clustering algorithms could enable us to differentiate MS from SAD and DAF [7, 8]. The current work implemented a hybrid ensemble that integrates a Graph Attention Network (GAT) for semi-supervised node classification [9] on a graph derived from tabular data with a Tabular Prior-Data Fitted Network (TabPFN) [10] for direct tabular classification (fig. 1). The GAT exploits the graph structure to propagate labels from a small, labeled subset to unlabeled nodes, while TabPFN serves as a strong baseline by approximating Bayesian inference [11] on tabular features without explicit graph modeling. The ensemble fuses their outputs to leverage both relational (graph-based) and feature-interaction (tabular prior-based) strengths, making it particularly effective for small datasets with categorical labels (i.e. MS, SAD and DAF entities).

**Fig. 1.**
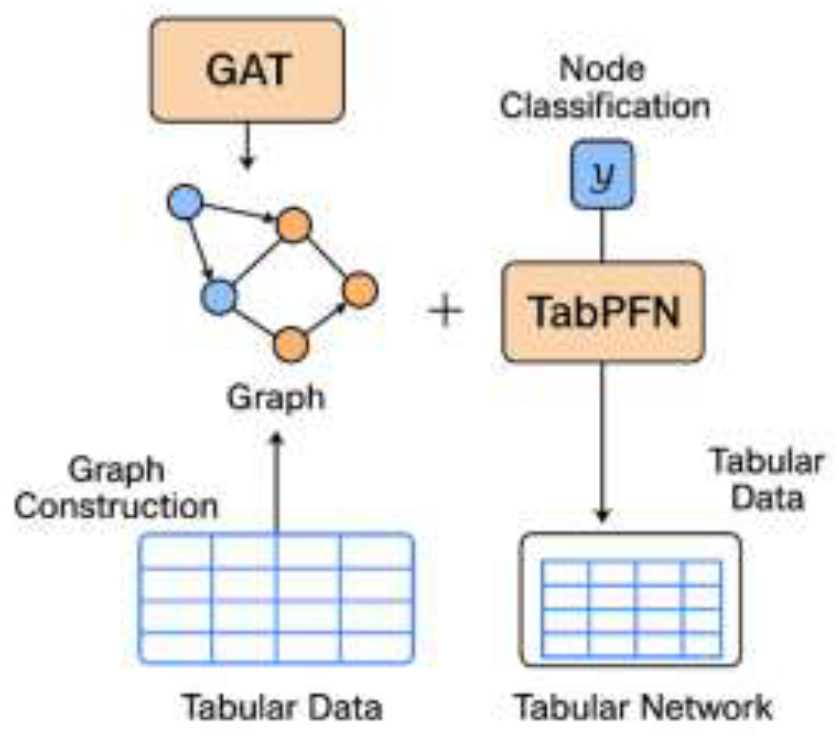
A visual representation of a hybrid ensemble of a GAT with a TabPFN.

## 2 Methodology

### 2.1 Data processing and feature selection

The tabular dataset was defined as 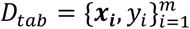, where ***x_i_*** ∈ *R*^*d*^ are the 240 input features, *y_i_* ∈ {0,1,2} are the categorical labels and *m=*396 patients. The missing entries were replaced with the column mean, and all features were standardized to mean 0 and variance 1 for numerical stability, 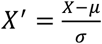 where *μ*, *σ* ∈ *R*^*d*^ are column wise mean and standard deviation. A random forest classifier was trained on the labeled training data {***x_i_***, *y_i_*|*i* ∈ *I*_*train*_} (40% stratified split). The Gini-based random forest importance of each feature was calculated as the average impurity decrease across all trees, 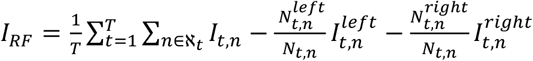, where 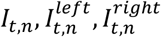 are the impurities at the parent and child nodes, *N*_*t,n*_ is the number of samples at node n in tree t, and 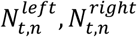 are the sample counts of the child nodes. The Gini impurity function in a K-classification problem can be defined as 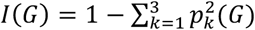 the class probability at a node G. The significance of each feature was assessed by calculating the permutation importance using the accuracy as a performance metric via paired t-test p-value. The top 50 features were selected based on Gini importance, with permutation means and t-test p-values printed for analysis [12].

### 2.2 Graph Attention Network (GAT)

GAT is a neural network architecture for graph-structured data, which estimates attention scores for each neighboring node to determine the importance of each neighborhood’s contribution. This differs from other approaches, like graph convolutional networks that use uniform or normalized weights.

An undirected graph *G =* (*V, E*) was initially constructed with a vertex set *V =* {1,2,3, …, *m*} and an edge set *E* determined by k-nearest neighbor relations (k=10). An initial adjacency was defined via k-NN (*k*=10) on full features using Euclidean distance. It was then replaced by thresholding Euclidean distances on training features only at the 80th percentile: *A*_*redefined,ij*_ = 1, if *d*_*ij*_ < *percentile*(*D*_*train*_, 80), where *D*_*train*_ are pairwise distances among training nodes (yielding a training-subgraph adjacency). Edge masks were used to mark which the nodes for training *M*_*train*_ (60%), *M*_*val*_ validation (20%) and *M*_*test*_ testing (20%).

GAT is a type of Graph Neural Network introduced by Veličković et al. (2018) [13], which can be described as a 2-layer attention mechanism [14] on *G*, allowing each node to treat neighborhood importance dynamically by computing attention coefficients that reflect the relative importance of each connection. Each node of GAT has a feature vector 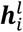 per layer, which is linearly transformed by a shared learnable weight matrix 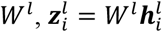 and used to compute an unormalized attention coefficients 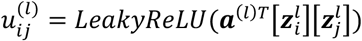 for each edge (i, j). ***a***^(*l*)^ is a learnable attention vector and [·] denotes concatenation. The attention coefficients have then been normalized 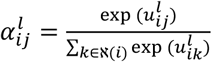, where ℵ(i) is the neighborhood of i. The node vector 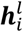 was then updated accordingly, 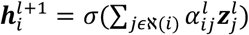, where *σ* is the exponential linear unit activation function (ELU).

Instead of having a single attention mechanism a multi-head (k=8) attention mechanism was employed to improve stability. For each node I,

- **Layer 1 (concatenate):** The edges weights were dynamically computed to modulate the importance of edges in the message passing process *GATConv* [15]. The weights *W*_*adj*_ are computed based on node features using a learnable projection, *ω_ij_* = *sigmoid*(*W*_*adj*_[***x**_i_*][***x***_*j*_]) [16], and are used to adjust the attention coefficients in the *GATConv* layer. Per head 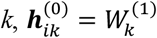 the attention coefficients are calculated as following:

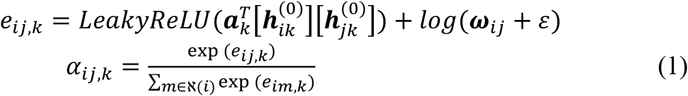

The node was then aggregated 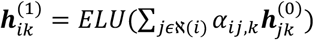, and concatenated 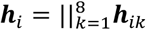

To prevent over-fitting, dropout with probability 0.5 was applied to 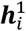.

- **Layer 2**: The second layer transforms node embeddings and aggregates information from neighboring nodes using a single attention head, 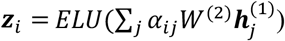

The loss function used for training was

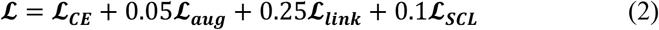

A cross entropy loss ***ℒ*_*CE*_**, was used to measure the difference between the predicted probability distribution *p*_*ic*_ *= softmax*(*z*_*ic*_) and the true class label *y_i_* ∈ {1,2,3} in the identified labeled nodes *I*_*train*_ used for training.

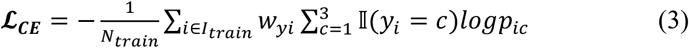

*w_yi_* is the class weight.

Augmented views were generated using feature dropout (p=0.1) and edge dropout to encourage consistency by introducing the augmentation loss, *ℒ*_***aug***_ = ***ℒ***_*CE*_(***y, z***_***aug***_).

A self-supervised contrastive loss *ℒ*_***SCL***_ only on the labeled training nodes was used to promote similar embeddings (of the first layer, 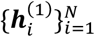) for nodes with the same label. This encourages the model to learn discriminative embeddings based on label similarity, without requiring additional augmentations or views beyond the existing graph structure. Each embedding is normalized, 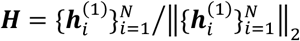 and a pairwise cosine similarity matrix is estimated *S*_*c*_ *=* ***H***^***T***^***H***/*τ*, scaled by temperature *τ* = 0.07. Note that unlike certain graph contrastive learning methods that create two augmented views of the graph, a single view was used, defining positive based on label equality. The exponential of S_*c*_ is then masked, using a positive pair mask *M* based on groundtruth labels *E = exp*(*S*_*c*_) ∗ *M*.

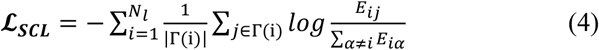

Γ(i) *=* {j: *y*_*j*_ = *y*_i_} (including self, *j*=*i*) are the same class labeled nodes and *N*_*l*_ is the number of labeled nodes.

A link prediction loss function *ℒ*_***link***_ is then used to predict whether an edge (*i, j*) *∈ E* (link) exists (positive, *E*_*pos*_) or not (negative, *E*_*neg*_) between two nodes in a graph. This encourages the embeddings ***h***_i_ to capture structural information, improving generalization in the semi-supervised setting. The loss function *ϕ*_*BCE*_, used was a binary cross entropy.

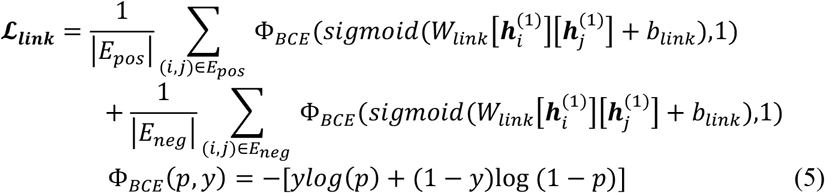

GATs loss function can be minimized using variants of stochastic gradient descent algorithms [17], with the Adam optimizer [18] being chosen, with a weight decay parameter 5e-4 and a learning rate 0.001.

Self-training (SST) [19] with entropy regularization [20] was performed after minimizing the cost function to leverage high-confidence predictions on unlabeled nodes and further improve the model’s performance in a semi-supervised setting. The inclusion of entropy regularization during the fine-tuning phase of self-training helps ensure that the model remains confident and generalizes well when incorporating pseudolabeled data. Initially the predicted probability that node *i* belongs to class *c* is derived from the softmax output of ***z***_*i,c*_, *p*_i,*c*_ *= softmax*(***z***_*i,c*_). In the self-training phase only unlabeled nodes with confidence *c*_*i*_ > 0.8 were considered, where *c*_*i*_ *= max*_*c*_*p*_*i,c*_. In each of 2 rounds, the top 50% (by *γ*=0.5) highest-confidence nodes among those >0.8 were pseudo-labeled via 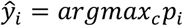, added to training, and fine-tuned for 50 epochs. An entropy term 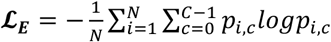 is then added to the fine tuning loss function, *ℒ*_***ft***_ *= ℒ*_*CE*_ + 0.1*ℒ*_***E***_.

### 2.3 Hybrid ensemble with TabPFN

TabPFN is a pretrained Bayesian meta-model that approximates full Bayesian inference *p*(*y*_*test*_|*x*_*test*_, *D*_*tab*_) *= ∫ p*(*x*_*test*_|*y*_*test*_, *φ*)*p*(*φ*|*D*_*tab*_)*d*φ for small tabular datasets, fitted on the (post-SST) labeled training subset of *D*_*tab*_, where parameters φ is typically a neural network. TabPFN approximates the mapping by training a transformer *ξ_θ_*, *ξ_θ_*(*y*_*test*_|*x*_*test*_, *D*_*tab*_) ≈ *p*(*y*_*test*_|*x*_*test*_, *D*_*tab*_). The predicted class probabilities are approximated by *p*_*tabPFN*_ *= softmax*(*ξ_θ_*(*y*_*test*_|*x*_*test*_, *D*_*tab*_)). Given the GAT output probabilities *p*_*GAT*_ *= softmax*(*z*_*c*_(*x*_*test*_)), the hybrid ensemble log-probabilities were formulated as: log *p*_*ens*_ *= αlogp*_*GAT*_ + (1 − *α*)*log*(*p*_*tabPFN*_ + 1*e* − 8) with *α =* 0.6. The ensemble predictions *y*_*ens*_, which combine outputs from the Graph Attention Network (GAT) and TabPFN classifier are given by *y*_*ens*_ *= argmax*_*c*_*p*_*ensi,c*_.

### 2.4 Statistical Analysis

The performance of the hybrid ensemble model was compared with multinomial logistic regression [21], where the probability of belonging to category k is obtained via the softmax function:

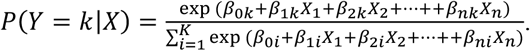

Its key assumptions are that observations are independent and there is no significant multicollinearity among predictors.

Both models were trained on 80% of the data and were validated on the excluded 20% testing dataset. Multiclass confusion matrices were used to evaluate the classification performance of the multinomial logistic regression and the proposed hybrid ensemble model. The row are the true labels of the classes, and the columns are the predicted ones. The diagonal elements of the matrix are the true positives of each class, and the off-diagonal elements are the misclassifications. Different metrics such as accuracy, precision, recall, and F1 score were used to evaluate the performance of the models. Accuracy measures the percentage of total predictions that were correct. Precision measures the percentage of the predicted positives that were true positives. Recall (sensitivity) measures the percentage of true positives that were correctly identified. The F1 score provides a single, balanced measure of a model’s performance by calculating the harmonic mean of precision and recall, which is especially useful when dealing with imbalanced datasets where accuracy alone might be misleading.

The metrics were reported for the 20% of the testing dataset, that was not included in the training of the classification models.

## 3 Results

All the features used in training according to the Gini importance score are shown in table 1. The addition of more features did not significantly improve the testing accuracy. The confusion matrix, accuracy, precision, recall, and F1-Score values were calculated and presented in Tables 2 and 3.

**Table 1.** Feature importance according to the Gini importance score generated by the Random Forest classifier.

| MS vs SAD vs DAF |  |
| --- | --- |
| CSF IgG | 0.044 |
| CSF glucose | 0.039 |
| Gamma globulins | 0.036 |
| Hgb | 0.036 |
| Vit-D | 0.035 |
| Age at 1st neurological symptom | 0.034 |
| Age | 0.032 |
| CSF white cells | 0.031 |
| CSF red cells | 0.030 |
| Brainstem signs | 0.030 |

**Table 2.** Confusion matrices of the multinomial logistic regression and the hybrid ensemble models for the classification of MS, SAD, DAF.

|  |  | Multinomial Logistic Regression |  |  | Hybrid ensemble model |  |  |
| --- | --- | --- | --- | --- | --- | --- | --- |
|  |  | Predicted labels |  |  | Predicted labels |  |  |
|  |  | MS | SAD | DAF | MS | SAD | DAF |
| True labels | MS | 39 | 1 | 10 | 49 | 0 | 0 |
|  | SAD | 0 | 2 | 4 | 1 | 3 | 2 |
|  | DAF | 18 | 0 | 6 | 13 | 4 | 7 |

**Table 3.** Evaluation of the multinomial logistic regression and the hybrid ensemble models in terms of accuracy, precision, recall, and F1-score for the classification of MS, SAD, DAF.

|  | Accuracy | F1-score | Precision | Recall |
| --- | --- | --- | --- | --- |
| Multinomial Logistic Regression | 0.588 | 0.482 | 0.550 | 0.454 |
| Hybrid ensemble model | 0.738 | 0.573 | 0.644 | 0.591 |

## 4 Conclusion

Systemic autoimmune diseases (SADs) may present with CNS demyelination, mimicking multiple sclerosis (MS), though driven by distinct immunopathogenic mechanisms. While ANA and aPL are commonly reported in MS, the role of specific SAD-related autoantibodies (ANA profile) in CNS demyelination remains unclear. The aims of this study were to overcome these diagnostic challenges, by investigating novel clustering algorithms that could differentiate MS from SAD and DAF.

This work implemented a hybrid model for semi-supervised tabular classification, integrating:

i. a Graph Attention Network with dynamic graph learning via Random Forest proximity and k-NN graphs and
ii. a TabPFN ensemble for improved predictions.

The model also performed feature selection, self-supervised contrastive learning, self-training and data augmentation. The achieved accuracy was 74%, which was significantly higher from the multinomial logistic regression (59%).

The results demonstrate the hybrid ensemble model effectiveness (74% accuracy) over logistic regression (59%), leveraging graph-based propagation, and ensemble with TabPFN for better differentiation of MS, SAD, and DAF.

Table 1 importance features include demographic factors like age and age at first neurological symptom (younger in MS vs. older in SAD/DAF); brainstem signs (more focal in MS); laboratory markers such as CSF IgG (elevated in MS), CSF glucose, gamma globulins, Hgb (lower in SAD related anemia), Vit-D (deficient in systemic autoimmunity), CSF white cells (mild elevation in active inflammation), and CSF red cells. These parameters underscore the model’s focus on biomarkers that capture organ specific CNS demyelination in MS versus systemic autoimmunity in SAD/DAF, aiding early differentiation and reducing misdiagnosis risks.

## Data Availability

All data produced in the present study are available upon reasonable request to the authors

## 5 Acknowledgements

This work was funded by the Sectoral Development Program (SDP 5223471) of the Ministry of Education, Religious Affairs and Sports, through the National Development Program (NDP) 2021-25.

